# Greater Mental Health Benefits Following Contemplative-Based Social Resilience Training Among Young Adults with Early-Life Adversity

**DOI:** 10.64898/2026.07.21.26358618

**Authors:** Aaron M. Eisen, Philippe Goldin, Jyoti Mishra, Elena Fromer, Lucy Kho, Aric A. Prather, Elissa S. Epel, the UC Climate Resilience Consortium

**Author notes:** Corresponding Author: Aaron M. Eisen.

## Abstract

Emerging evidence suggests that adults with a history of early life adversity (ELA), while more susceptible to psychopathology, may also be particularly sensitive to the benefits of contemplative practices. In the present study, we examined whether ELA was associated with greater mental health benefits following a contemplative-based social resilience training program, administered as a university elective course across all ten campuses of the University of California (*n* = 321; median age = 21 years; 74% female). While significant improvements in mental health were observed for all participants, those with higher ELA exhibited greater reductions in mental distress (3.5-fold larger, *p* = .007) and greater increases in well-being (2.5-fold larger, *p* = .010) relative to those with lower ELA. Replication in future studies and further research on the mechanisms underlying this enhanced benefit may improve our understanding of how adults with a history of ELA recover and may ultimately thrive.

## Background

Over two decades of research has consistently documented that chronic exposure to early life adversity (ELA), whether abuse, maltreatment, or household dysfunction, is a potent social determinant of negative health outcomes across the lifespan (Agorastos et al., 2018, 2019; Berens et al., 2017; Danese & Lewis, 2017; Felitti et al., 1998; Malave et al., 2022; Taylor, 2010; Taylor et al., 2011), with especially pervasive effects on mental health and well-being (McLaughlin et al., 2020; Nelson et al., 2025). It is well-established that ELA at least doubles the risk of a broad array of psychiatric comorbidities during adulthood, including depressive and anxiety disorders, internalizing disorders, post-traumatic stress disorder, and suicidality (Abate et al., 2025; Annor et al., 2026; Sahle et al., 2022; Thurston et al., 2025). Unfortunately, ELA is very common, with recent estimates suggesting that over one-third of the United States adult population has experienced maltreatment during childhood (Swedo et al., 2023), with a rising prevalence (Kumar et al., 2024). The Centers for Disease Control and Prevention (CDC) recognizes these associations as a preventable public health crisis and has estimated that ELA may account for up to 44% of the national prevalence of depressive disorders (Gervin et al., 2022; Jones et al., 2020). Hence, there is an urgent need for interventions to mitigate the lifelong consequences of ELA.

While there is growing interest in how ELA can affect treatment outcomes within intervention contexts, the literature to date has been mixed. Several seminal studies have shown that traditional interventions, including psychotherapy and psychopharmacology approaches, are less effective at treating depressive disorders among adults with ELA versus those without such history (Dos Santos et al., 2026; Nemeroff et al., 2003; Nanni et al., 2012; Williams et al., 2016), while other studies have found no differences in treatment outcomes (Childhood Trauma Meta-Analysis Study Group, 2022; Ferensztajn-Rochowiak et al., 2026; Lewis et al., 2010; Medeiros et al., 2021). On the other hand, several studies have shown that process-oriented psychosocial interventions, including psychodynamic and emotion-focused therapy, are more effective at reducing depressive disorders among adults with ELA compared to those without such history (Aardal et al., 2026; Goerigk et al., 2024; Heinonen et al., 2018; Krakau et al., 2024).

Importantly, beyond patients with clinical depressive disorders, mindfulness-based interventions (MBI) are contemplative approaches that show considerable promise in reducing mental distress among adults with ELA. In a scoping review of 17 studies, Joss and Teicher (2021) highlighted the clinical effectiveness of mindfulness meditation training in reducing psychological symptoms among adults with ELA, with significant reductions in stress, anxiety, depression, substance use, and post-traumatic stress across various MBIs (e.g., mindfulness-based stress reduction [MBSR], mindfulness-based cognitive therapy [MBCT], mindfulness-based relapse prevention [MBRP], and mindful self-compassion [MSC]). In a recent randomized controlled trial, Joss et al. (2024) found that among young adults with ELA participating in an 8-week MBI, increases in mindfulness predicted reductions in stress, somatization, and hostility scores, while this pattern was not observed among young adults with ELA participating in a stress management education (control) condition (without mindfulness training). In a systematic review of 13 mixed methods studies, Moyes et al. (2022) found that various forms of mindfulness training were effective in reducing mental distress, negative moods, and anxiety among adults with ELA, with qualitative themes suggesting that increases in psychological flexibility, management of emotions, social skills, and coping strategies were instrumental for these reductions.

Several studies have also found that adults with ELA exhibit greater benefits from mindfulness training compared to those without such history. In a randomized controlled trial, Andersen et al. (2021) found that women with ELA who participated in an MBSR intervention exhibited specific improvements in sensitization-related outcomes, reduced cortisol responses to an experimental stressor (Trier Social Stress Test), and greater benefits in describing emotions and impulse control relative to women without ELA, and relative to women in the social support (control) condition (without mindfulness training). In a randomized controlled trial of MBCT, Williams et al. (2014) found that ELA significantly moderated treatment outcomes, such that significant reductions in risk of depression relapse between MBCT versus treatment-as-usual (TAU) were only observed for adults above the median severity level for ELA.

Put together, this evidence suggests that mindfulness-based approaches are a feasible alternative for traditional interventions that show considerable promise in reducing mental distress among adults with ELA. Specifically, by cultivating present-moment awareness, non-judgement, and self-compassion, mindfulness training can help to reduce rumination and worries, improving coping with negative cognitions such as self-blame, shame, and guilt, and support overcoming low self-esteem, which are especially prevalent within this population and major contributing factors for psychopathology (Joss & Teicher, 2021).

### Present Study

However, mindfulness approaches are just one contemplative practice within a broader resilience framework – including building interconnection to the natural world through experiential practices, cultivating worldviews based on interdependence through pedagogical approaches, and strengthening social resilience through reflective dialogue, values clarification, cross-cultural communication, and psychological flexibility to promote prosocial behavior and collective actions (Epel et al., 2025). Although contemplative practices beyond mindfulness are relatively understudied among adults with ELA (Scafuto et al., 2025), integrating multiple components of this broader resilience framework into a single approach may provide complementary pathways for strengthening individual and collective resilience.

One particularly promising example is the integration of mindfulness with the therapeutic potential of nature (for an excellent review on the health benefits of nature contact, see Frumkin et al., 2017). Growing evidence suggests that nature-based practices buffer against many of the mental health risks associated with ELA (Denker & Faber Taylor, 2026; Touloumakos & Barrable, 2020; White et al., 2023), while emerging evidence indicates that mindfulness practices in natural environments may produce enhanced benefits, while also cultivating nature connection and ecological awareness (Vecchio et al., 2026). A recent global analysis across 75 countries also demonstrated robust evidence for an association between nature connection, mindfulness, and multiple aspects of well-being, including purpose in life, hope and optimism, and life satisfaction (Barbett et al., 2026).

By contrast, the majority of research on traditional interventions has focused on reducing negative symptoms among adults with ELA, and further research is needed to better understand how to foster positive emotions and well-being in this population. This is critical because even if adults with ELA do not decrease to low levels of distress, they can still develop well-being given supportive contexts, helping to improve their entire mental health continuum (Keyes, 2002). Taken together, these gaps underscore the need to evaluate multi-component contemplative-based and social resilience approaches among adults with ELA and to examine changes across both negative and positive domains of the mental health continuum.

Hence, the objective of the present study was to examine whether ELA may be associated with greater mental health benefits from a contemplative-based social resilience (CBSR) course, a structured training program championing the components of this broader resilience framework to reduce eco-anxiety and improve mental health and well-being in university students (Epel et al., 2025). Specifically, we examined the moderation effect of ELA (experiences of abuse and neglect before age 18) on changes in mental distress (symptoms of stress, anxiety, and depression) and mental well-being (perceptions of emotional, social, and psychological well-being) pre-to-post CBSR course. Based on prior research, we hypothesized that while all participants would exhibit mental health benefits, those with ELA would exhibit even more pronounced benefits, as exemplified by greater decreases in mental distress and greater increases in mental well-being before versus after the course, relative to those without a history of ELA.

## Method

### Participants

Participants included 461 university students enrolled in a for-credit contemplative-based social resilience course across all ten campuses of the University of California (UC) academic system during the Spring quarters of 2024 (*n* = 169) and 2025 (*n* = 292). Students were informed of the course through listservs, newspaper articles, online course announcements, academic advisors, and word of mouth. There were no exclusion criteria. Students who agreed to participate in this study provided informed consent, and measurements were obtained from a self-report battery administered online using Qualtrics. Data were deidentified, and this study received ethical approval from the University of California, San Diego (#180140) and the University of California, San Francisco (#24-42083).

### Contemplative-Based Social Resilience Course

The CBSR course was delivered in-person over a quarter or semester and facilitated by university faculty and experienced meditation instructors at each of the ten campuses. Guided by multiple frameworks, including Acceptance Commitment Training (ACT), Self-Determination Theory (SDT), and Bandura’s Social Learning Theory, each class consisted of contemplative practices, experiential learning, didactic content, and structured dialogues with selected content on climate science, ecological challenges, and psychosocial resilience (Epel et al., 2025).

Students began with guided mindfulness, compassion, and emotional-awareness practices designed to strengthen emotion regulation, distress tolerance, and psychological flexibility. These capacities were then extended through nature-based meditation, outdoor immersion, nature-based retreats, and other experiential exercises intended to cultivate an embodied sense of connection with the natural world. Didactic materials, expert perspectives from thought leaders, and facilitated inquiry invited students to consider the interdependence of self, others, and the broader living world, while values clarification, reflective dialogue, perspective-taking, and cross-cultural communication helped them relate these perspectives to their own lives and to the experiences of others. Small-group discussions and collaborative activities provided opportunities to practice generous listening, translate clarified values into prosocial behavior, and develop shared efficacy for collective action through collaborative climate-action projects.

Instructors supported this process by modeling mindfulness, compassion, openness, and psychological flexibility and by cultivating a psychologically safe environment in which students could share personal responses to course materials, explore their relevance to their lives, and engage in respectful, supportive dialogue.

### Study Measures

#### Early Life Adversity

Exposure to ELA was assessed at baseline using the well-validated 28–item version of the Childhood Trauma Questionnaire (CTQ–28; Bernstein et al., 2003). Subscales for emotional, physical, and sexual abuse, and emotional and physical neglect were calculated by reverse scoring and summing the corresponding items, which were then summed to calculate a total score for ELA. Internal consistency for the total score was excellent (α = 0.92) and satisfactory for the subscales (α = 0.72 – 0.90). Cutoff scores for moderate-severe severity are: +12 points for physical abuse, sexual abuse, and physical neglect; +15 points for emotional abuse; +17 points for emotional neglect (Bernstein & Fink, 1998); and +35 points for the total ELA score (consistent with prior literature, e.g., Singha et al., 2024). Scores for emotional, physical, and sexual abuse, and physical and emotional neglect were summed to create dimension scores for early life abuse (α = 0.90) and neglect (α = 0.85).

#### Mental Distress

Symptoms of mental distress were assessed at baseline and immediately following the CBSR course using the well-validated 21–item version of the Depression, Anxiety, Stress Scale (DASS–21; Lovibond & Lovibond, 1995). Subscales for stress, anxiety, and depression were calculated by summing the corresponding items which were multiplied by a factor of 2. Subscale scores were then summed to calculate a total score for mental distress. Internal consistency for the total score was excellent (α = 0.93) and satisfactory for the subscales (α = 0.80 – 0.88). Cutoff scores for moderate and severe severity are: +19 and +26 points for stress; +10 and +15 points for anxiety; and +14 and +21 points for depression, respectively (Lovibond & Lovibond, 1995).

#### Mental Well-Being

Perceptions of mental well-being (“flourishing”) were assessed at baseline and immediately following the CBSR course using the well-validated Mental Health Continuum Short Form (MHC–SF; Keyes, 2005). Subscales for perceptions of emotional (hedonic), social (eudaimonic), and psychological (eudaimonic) well-being were calculated by summing the corresponding items, which were then summed to calculate a total score for mental well-being. Internal consistency for the total score was excellent (α = 0.91) and satisfactory for the subscales (α = 0.79 – 0.87). There are no cutoffs for the total score; higher values indicate greater mental well-being.

#### Analytical Approach

As our analytical approach required complete data for model covariates, participants missing ELA data (*n* = 131; 28.4%) and demographic data (*n* = 8; 1.7%) were excluded, along with one participant who did not provide mental health data at any time point. Hence, the final analytical sample included a total of 321 participants, and no significant differences were observed between participants missing data and the analytical sample across our study variables (Table S1).

Our hypothesis was evaluated using linear mixed effects models with restricted maximum likelihood estimation. Each model contained random intercepts and a two-way interaction term (ELA x Time) to examine the moderation effect of ELA on changes in mental distress (DASS-21) and well-being (MHC-SF) pre-to-post CBSR course. Interactions were probed using simple slopes (pre and post CBSR course) and marginal means (±1 *SD*) with pairwise contrasts. Standardized effect sizes for within-person changes (Cohen’s *d*) were calculated using the baseline standard deviation to account for baseline variability. Models were adjusted for years of age and gender identity. ELA was mean-centered and variance inflation factor values provided no evidence of multicollinearity (cutoff value > 2.0). Modeling assumptions were affirmed and model fit was assessed prior to interpreting our results.

Specificity across domains of mental distress (symptoms of stress, anxiety, and depression) and well-being (perceptions of emotional, social, and psychological well-being) was examined using multivariate mixed effects models with a three-way interaction term (ELA x Time x Outcome Subscale). Lower order terms were modeled and correlated random intercepts were specified (within-person) for each subscale. Residuals were allowed to vary by subscale and correlate freely (within-person) at each time point. Secondarily, the unique contribution of early life abuse (emotional, physical, and sexual) versus neglect (emotional and physical) was examined by specifying interaction terms for both dimensions within each model. Differences in the moderation effect between these terms were determined using a Wald (χ²) test.

Although we used alphas (0.05 level) to report significant results, our interpretations focused on effect sizes and confidence intervals across all models, regardless of statistical significance. Data were analyzed using R (Version 4.4.3).

## Results

The analytical sample consisted of 321 participants who were primarily female (73.8%) undergraduate students (83.0%) with a high representation of Asian (30.3%), Hispanic (28.1%), and White (16.9%) young adults (Median = 21.0 years, IQR = 20.0 – 23.0 years; Table 1). ELA was remarkably prevalent within the sample, with 60.4% (*n* = 194) reporting moderate to severe exposure across subdomains (CTQ > 35 points). Emotional abuse was the most common ELA (*n* = 100; 31.2%), followed by physical neglect (*n* = 81; 25.2%), sexual abuse (*n* = 63; 19.6%), emotional neglect (*n* = 57; 17.8%), and physical abuse (*n* = 49; 15.3%). In terms of demographics, ELA was less common among White participants, and participants who endorsed non-cisgender identities trended toward higher exposure (Table 1). Bivariate correlations across our study variables at baseline and across the different subdomains of ELA are provided in the supplemental materials (Figures S1-S2).

**Table 1.** Participant Characteristics by ELA Status.

| Characteristic | N (%) |  |  | p-value<sup>1</sup> |
| --- | --- | --- | --- | --- |
|  | Total Sample<br>(n = 321) | Low ELA<br>(n = 127) | High ELA<br>(n = 194) |  |
| Years of Age |  |  |  |  |
| Below 25 | 258 (80.4%) | 100 (78.7%) | 158 (81.4%) | p = .792 |
| 25 to 30 | 29 (9.0%) | 12 (9.4%) | 17 (8.8%) |  |
| Above 30 | 34 (10.6%) | 15 (11.8%) | 19 (9.8%) |  |
| Gender Identity |  |  |  |  |
| Cis-Female | 237 (73.8%) | 101 (79.5%) | 136 (70.1%) | p = .169 |
| Cis-Male | 63 (19.6%) | 21 (16.5%) | 42 (21.6%) |  |
| Other | 21 (6.5%) | 5 (3.9%) | 16 (8.2%) |  |
| Race & Ethnicity |  |  |  |  |
| Asian | 97 (30.8%) | 36 (28.3%) | 61 (31.4%) | p = .004<sup>2</sup> |
| Hispanic | 90 (28.6%) | 27 (21.3%) | 63 (32.5%) |  |
| White | 54 (17.1%) | 32 (25.2%) | 22 (11.3%) |  |
| Mixed | 46 (14.6%) | 20 (15.7%) | 26 (13.4%) |  |
| Black | 19 (6.0%) | 3 (2.4%) | 16 (8.2%) |  |
| Other | 9 (2.9%) | 4 (3.1%) | 5 (2.6%) |  |
| Student Status |  |  |  |  |
| Undergrad | 264 (83.0%) | 102 (80.3%) | 162 (83.5%) | p = .818 |
| Graduate | 33 (10.4%) | 14 (11.0%) | 19 (9.8%) |  |
| Post-Degree | 12 (3.8%) | 6 (4.7%) | 6 (3.1%) |  |
| Other | 9 (2.8%) | 4 (3.1%) | 5 (2.6%) |  |

<sup>1</sup> Fisher's exact test between low versus high ELA groups (CTQ > 35 points).
<sup>2</sup> Non-significant when excluding the "White" category ( $p = .184$ ).

### Mental Distress

In the overall sample, there were significant reductions in mental distress symptoms from pre-to-post CBSR course (*M*Δ = −5.94, *d* = 0.25, *p* < .001). Interaction effects indicated that ELA was associated with greater symptom reductions over time (*B* = −0.234, *p* = .007). Marginal means revealed that reductions in mental distress were 3.5-fold greater for participants with high adversity (+1 *SD*: *M*Δ = −9.29, *d* = 0.39, *p* < .001) versus low adversity (−1 *SD*: *M*Δ = −2.65, *d* = 0.11, *p* = .132), while simple slopes showed that the association between ELA and greater mental distress was weaker pre-to-post CBSR course (*B* = 0.544, *p* < .001 vs *B* = 0.310, *p* < .001). The multivariate model did not provide evidence of domain specificity (*F*(2,1340) = 0.11, *p* = .894), suggesting that the moderation effect of ELA (greater symptom reductions over time) was consistent across domains of stress (*B* = −0.083, *p* = .022), anxiety (*B* = −0.095, *p* = .002), and depression (*B* = −0.093, *p* = .007; Figure 1; Table S2). No significant differences were observed between early life abuse and neglect (χ²(1) = 1.14, *p* = .285).

**Figure 1.**
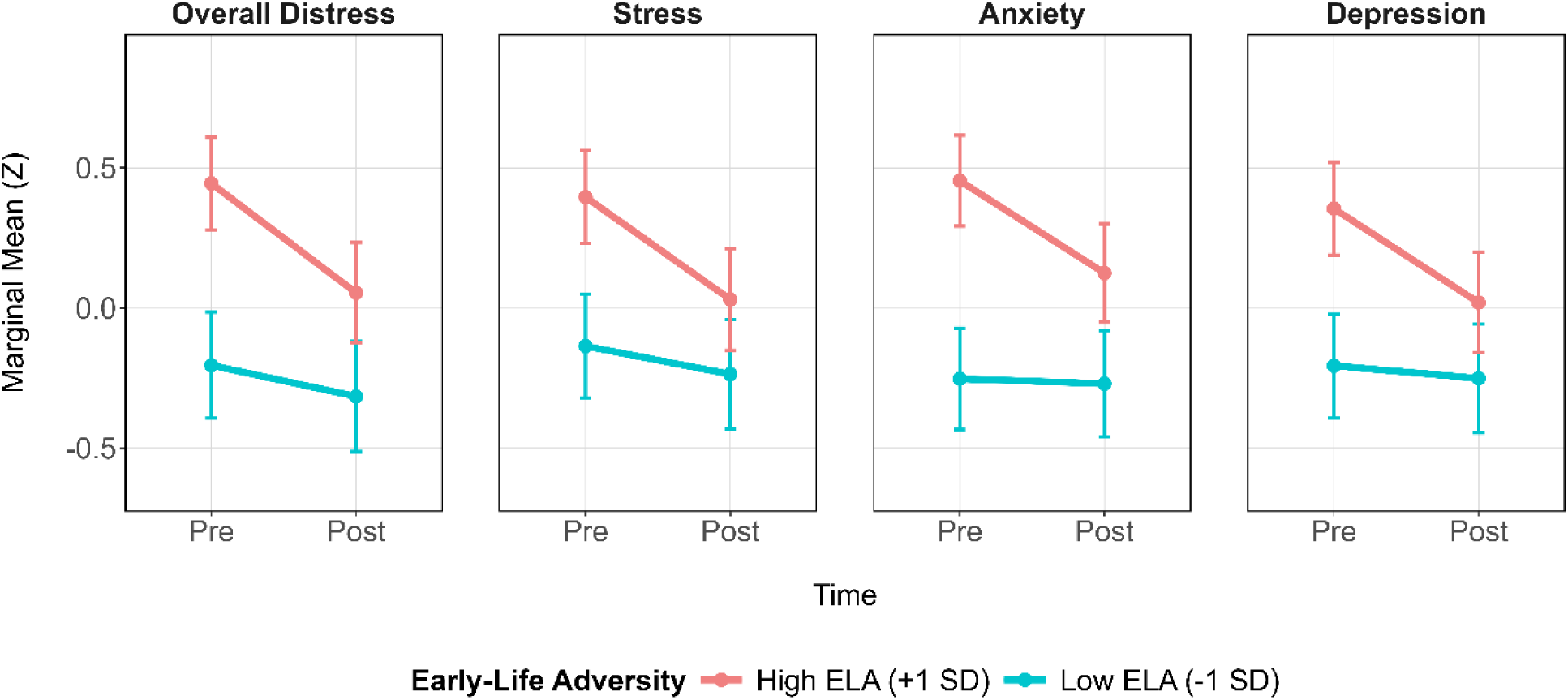
Reductions in Mental Distress by ELA Status *Note.* Estimated marginal means (z-scored) from the mixed effects models visualizing mental distress (overall and subscales) pre-to-post CBSR course, stratified by high (+1 *SD*) versus low (−1 *SD*) ELA. Overall mental distress estimates are from the linear mixed effects model, while subscale estimates are from the multivariate mixed effects model. Models were adjusted for age and gender identity. Error bars represent 95% confidence intervals.

### Mental Well-Being

In the overall sample, there were significant increases in mental well-being from pre-to-post CBSR course (*M*Δ = 4.47, *d =* 0.37, *p* < .001). Interaction effects indicated that ELA was associated with greater improvements in mental well-being over time (*B* = 0.135, *p* = .010). Marginal means revealed that improvements in well-being were 2.5-fold greater for participants with high adversity (+1 *SD*: *M*Δ = 6.38, *d* = 0.52, *p* < .001) versus low adversity (−1 *SD*: *M*Δ = 2.53, *d* = 0.21, *p* = .016), while simple slopes showed that the association between ELA and lower mental well-being was weaker pre-to-post CBSR course (*B* = −0.261, *p* < .001 vs *B* = −0.126, *p* = .037). The multivariate model did not indicate domain specificity (*F*(2,1073) = 1.70, *p* = .184) suggesting that the moderation effect of ELA (greater increases in well-being over time) was consistent across domains of emotional (*B =* 0.024, *p* = .028), social (*B* = 0.064, *p* = .010), and psychological well-being (*B* = 0.052, *p* = .050; Figure 2; Table S3). No significant differences were observed between childhood abuse and neglect (χ²(1) = 1.24, *p* = .266).

**Figure 2.**
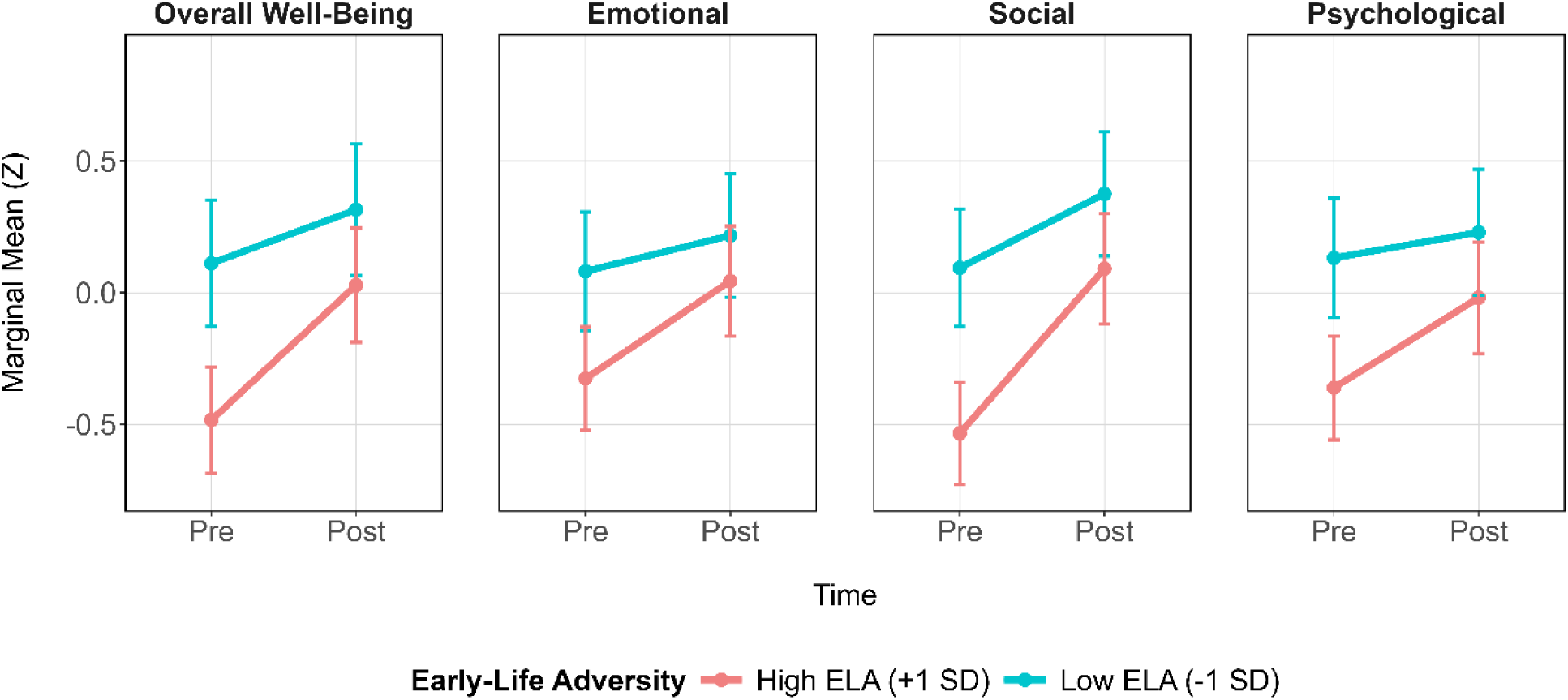
Increases in Mental Well-Being by ELA Status *Note.* Estimated marginal means (z-scored) from the mixed effects models visualizing mental well-being (overall and subscales) pre-to-post CBSR course, stratified by high (+1 *SD*) versus low (−1 *SD*) ELA. Overall well-being estimates are from the linear mixed effects model, while subscale estimates are from the multivariate mixed effects model. Models were adjusted for age and gender identity. Error bars represent 95% confidence intervals.

### Mental Distress and Well-Being

To examine whether the moderation effect of ELA differed between improvements in mental distress and well-being pre-to-post CBSR course, we fit another multivariate mixed effects model with a three-way interaction term (ELA x Time x Mental Health Domain), specifying overall scores for mental distress and well-being as correlated outcomes. Results from this model were consistent with the primary (independent) mixed effects models (Table S4), and the three-way interaction term was significant (*F*(1,660) = 11.52, *p* < .001) indicating that ELA was more strongly associated with reductions in mental distress (*M*Δ = −2.28 vs −9.11, *p* = .006; 4.0-fold greater reductions) versus increases in well-being (*M*Δ = 2.68 vs 6.31, *p* = .013; 2.4-fold greater increases) pre-to-post CBSR course.

## Discussion

In the present study, we examined the mental health benefits of a contemplative-based social resilience (CBSR) course, a structured training program designed to reduce eco-anxiety and improve mental health and well-being in university students across multiple university campuses. Consistent with our hypothesis, ELA significantly moderated changes in mental distress and well-being pre-to-post CBSR course. Although significant improvements in mental health were observed for all students, those with ELA exhibited significantly greater reductions in mental distress and increases in well-being following the CBSR course (2.4-fold to 4.0-fold larger), as compared to participants without such history. ELA was still associated with worse mental health following the CBSR course, but this association was noticeably attenuated relative to baseline and, in some cases, no longer significant.

### Theoretical Implications

Considering that ELA was strongly associated with greater mental distress and lower well-being at baseline, one possible explanation for these findings is that participants with ELA simply had “more room to improve” from the supportive context created by CBSR. This interpretation aligns with evidence that mindfulness training may positively impact the same neurobiological pathways through which ELA becomes biologically embedded (e.g., neural networks involved in self-regulation, immune signaling and inflammation, telomere biology, epigenetic alterations), helping to mitigate the risk of psychopathology over the life course (for a review, see Sun et al., 2022). In support of this idea, a recent mechanistic study of mindfulness training among adults with opioid use disorder found a distinct mechanistic pathway for symptom improvements among participants with ELA, specifically reductions in self-critical rumination, while this was not observed among participants with ELA in the recovery support (control) group (Joss et al., 2025).

However, another non-exclusive explanation is that adults with ELA are more sensitive to both the risks and benefits of their environmental conditions in a context-dependent manner (Differential Susceptibility; for a review, see Boyce, 2016). From this perspective, the same neurobiological pathways that induce susceptibility to stress and adversity might also confer enhanced responsivity to the supportive aspects of the environment, including the therapeutic potential of nature exposure (see Frumkin et al., 2017). Emerging evidence is consistent with this possibility. In a randomized controlled trial, Eisen et al. (2024) found that neurobiological correlates of ELA (inflammatory reactivity and glucocorticoid resistance; Nusslock & Miller, 2016) were associated with greater stress recovery following an acute stressor in an outdoor nature versus an indoor office setting. In a subsequent study, Eisen et al. (2026) found that adults with a history of ELA exhibited greater health benefits from residential nature exposure, as exemplified by lower levels of fasting blood glucose when living in greener neighborhoods.

Together, these perspectives suggest that the greater improvements observed among participants with ELA may reflect both greater baseline need and greater responsiveness to the supportive context created by CBSR. Rather than functioning as a single-component approach, CBSR integrates contemplative practice, nature exposure, social resilience building, and collective action, which may jointly provide opportunities for internal regulation, psychological restoration, and reconnection with others and the natural world (Epel et al., 2025). Thus, the present findings raise the possibility that approaches combining contemplative practices (such as mindfulness) and resilience components (such as nature connection) may be particularly well suited for supporting mental health and well-being among young adults with ELA.

### Research Implications

One of the most important implications of our findings is that participants with ELA not only exhibited greater reductions in mental distress, but also greater improvements in mental well-being following the CBSR course. This distinction is critical, as while current psychotherapy and psychopharmacological approaches are effective at reducing mental distress (Leichsenring et al., 2022), they are relatively ineffective at improving positive emotions and mental well-being (Craske et al., 2016, 2019, 2023; Dunn, 2012; Dunn et al., 2019). When conceptualizing distress and well-being as interrelated, but distinct domains of the broader mental health continuum (Keyes, 2005), it is clearer why focusing exclusively on distress limits both the clinical effectiveness of treatments and our scientific understanding of how individuals, especially those with a history of ELA, actually recover and may ultimately thrive (Belsky & Pluess, 2013; Ellis et al., 2011, 2017). Hence, we urge future investigators to measure the full range of the mental health continuum, as far too often, positive outcomes are treated as just the absence of disease or dysfunction rather than enhanced functioning or flourishing (Belsky & Pluess, 2009).

### Clinical Implications

The present findings suggest that CBSR may be useful as a campus-based approach to improve the mental health of university students, and especially those with a history of ELA. Even if the larger benefits observed in this study are due in part to a worse baseline level and more room for improvement, the exaggerated benefit is antithetical to the common belief of worse outcomes. It may be that the type of intervention determines whether there is greater benefit, with contemplative and nature-based approaches being particularly trauma sensitive and beneficial for ELA populations (Andersen et al., 2021; Denker & Faber Taylor, 2026; Joss & Teicher, 2021; Moyes et al., 2022; Scafuto et al., 2025; Touloumakos & Barrable, 2020; White et al., 2023).

### Strengths and Limitations

Notably, our study had several strengths, including the use of a large prospective multi-site sample, and our specification of variables across domains of the mental health continuum (mental distress and well-being) and dimensions of ELA (abuse and neglect) that were analyzed using multi-level and multi-variate modeling approaches.

At the same time, several limitations should be considered. The use of a non-randomized design without a control group limits causal inference that should be confirmed in future studies. Because our sample primarily consisted of female undergraduate students who were under the age of 30, additional research is needed to examine the generalizability of our findings to other populations. Future investigators are also encouraged to examine the influence of family dynamics on these associations, as highly supportive parental relationships have been shown to protect against ELA-induced psychopathology risk during adulthood (Farrell et al., 2017).

There was also a proportion of missing data for the ELA assessment (28.4%), as this measure was only offered during a supplemental post-course assessment for the 2024 cohort, but was administered at baseline for the 2025 cohort. Therefore, these data are considered to be missing completely at random, and no significant differences were observed between participants missing data and the analytical sample across our study variables (Table S1). Although we adjusted for years of age and gender identity, it is possible that other unassessed factors may have contributed to our findings (e.g., maternal relationships). Looking forward, future studies with more representative samples, rigorous assessments of other confounders, and randomized experimental designs to support causal modeling will be helpful.

## Conclusion

The present findings support CBSR as a promising, campus-based approach for promoting mental health among university students. Participation in the course was associated with reductions in mental distress and increases in mental well-being, with especially pronounced improvements among students with a history of ELA. By integrating contemplative practices, nature-based approaches, and social resilience building, CBSR may provide multiple complementary pathways for addressing the enduring mental health consequences of ELA and strengthening individual and collective resilience. Replication in future studies and further research on the mechanisms underlying this enhanced benefit may improve our understanding of how adults with a history of ELA recover and may ultimately thrive.

## Data Availability

All data produced in the present study are available upon reasonable request to the authors

## Acknowledgments

We wish to thank each instructor of the course who contributed their expertise, time, and passion. We also greatly appreciate each student who chose to take the course and participate in this study. UC Climate Resilience Consortium: Elissa Epel, Philippe Goldin, Jyoti Mishra, Gail Wright, Alison Holman, Cassandra Vieten, Larisa Castillo, Viveka Ramel, Diana Hill, Dana Garfin, Rebecca Peters, Anthony Maes, Anne-Marie Brest, Robert Lurye, Vickie Mays, Cindy Wong, Greg Crespo, and Amanda Frazier.

