## Supplemental Materials for "Greater Mental Health Benefits Following Contemplative-Based Social Resilience Training Among Young Adults with Early-Life Adversity"

### Table S1

*Participants Missing Data vs Analytical Sample Across Study Variables*

| Characteristic | *M* (± *SD*) or *N* (%) | | | p-value[1] |
|---|---|---|---|---|
| | Total Sample (*n* = 461) | Missing Data (*n* = 140) | Analytic Sample (*n* = 321) | |
| Years of Age | 23.0 (± 7.2) | 22.8 (± 7.3) | 23.0 (± 7.2) | $p = .814$ |
| Female Gender | 306 (72.5%) | 69 (68.3%) | 237 (73.8%) | $p = .293$ |
| Mental Distress | 38.0 (± 24.1) | 40.3 (± 27.9) | 37.4 (± 23.0%) | $p = .393$ |
| Mental Well-Being | 41.3 (± 12.2) | 40.6 (±13.3) | 41.4 (± 12.2) | $p = .833$ |

[1] T-test or $\chi^2$ test between participants missing data versus analytical sample at baseline.

**Table S2**

*Interaction Effects for ELA Moderating Mental Distress*

| | Marginal Means | | Simple Slopes | |
|---|---|---|---|---|
| | **Low ELA (-1 *SD*)** | **High ELA (+1 *SD*)** | **Baseline** | **Post-Course** |
| **Overall Distress** | *M*Δ = -2.65<br>(-6.09, 0.80)<br>*d* = 0.11 | *M*Δ = -9.29***<br>(-12.76, -5.82)<br>*d* = 0.39 | *B* = 0.544***<br>(0.375, 0.713) | *B* = 0.310***<br>(0.130, 0.490) |
| **Depression Subscale** | *M*Δ = -0.40<br>(-1.76, 0.95)<br>*d* = 0.04 | *M*Δ = -3.03***<br>(-4.39, -1.67)<br>*d* = 0.33 | *B* = 0.179***<br>(0.173, 0.245) | *B* = 0.087*<br>(0.016, 0.157) |
| **Anxiety Subscale** | *M*Δ = -0.15<br>(-1.37, 1.07)<br>*d* = 0.02 | *M*Δ = -2.84***<br>(-4.08, -1.60)<br>*d* = 0.33 | *B* = 0.214***<br>(0.153, 0.275) | *B* = 0.120***<br>(0.054, 0.185) |
| **Stress Subscale** | *M*Δ = -0.89<br>(-2.32, 0.54)<br>*d* = 0.10 | *M*Δ = -3.24***<br>(-4.68, -1.79)<br>*d* = 0.36 | *B* = 0.166***<br>(0.102, 0.231) | *B* = 0.084*<br>(0.014, 0.154) |

*Note.* Marginal means showing pre-to-post changes *M*Δ (95% CI) in overall mental distress and subscales at low (-1 *SD*) versus high (+1 *SD*) ELA. *B* (95% CI) reports unstandardized simple slopes and standardized mean differences (Cohen's *d*) were calculated using the baseline standard deviation of each outcome. Overall distress estimates are from the linear mixed effects model, while subscale estimates are from the multivariate mixed effects model. Models were adjusted for age and gender identity.

* $p < .05$, ** $p < .01$, *** $p < .001$

**Table S3**

*Interaction Effects for ELA Moderating Mental Well-Being*

| | Marginal Means | | Simple Slopes | |
|---|---|---|---|---|
| | **Low ELA (-1 *SD*)** | **High ELA (+1 *SD*)** | **Baseline** | **Post-Course** |
| **Overall Well-Being** | $M\Delta = 2.53$* (0.48, 4.59) $d = 0.21$ | $M\Delta = 6.38$*** (4.33, 8.42) $d = 0.52$ | $B = -0.261$*** (-0.371, -0.151) | $B = -0.126$* (-0.244, -0.008) |
| **Emotional Subscale** | $M\Delta = 0.40$ (-0.03, 0.82) $d = 0.14$ | $M\Delta = 1.07$*** (0.65, 1.50) $d = 0.38$ | $B = -0.044$*** (-0.070, -0.019) | $B = -0.021$ (-0.048, 0.007) |
| **Social Subscale** | $M\Delta = 1.49$** (0.52, 2.46) $d = 0.29$ | $M\Delta = 3.30$*** (2.33, 4.27) $d = 0.64$ | $B = -0.121$*** (-0.167, -0.076) | $B = -0.058$* (-0.107, -0.008) |
| **Psychological Subscale** | $M\Delta = 0.59$ (-0.44, 1.62) $d = 0.10$ | $M\Delta = 2.05$*** (1.02, 3.08) $d = 0.34$ | $B = -0.109$*** (-0.162, -0.056) | $B = -0.058$* (-0.114, -0.001) |

*Note.* Marginal means showing pre-to-post changes $M\Delta$ (95% CI) in overall mental well-being and subscales at low (-1 *SD*) versus high (+1 *SD*) ELA. *B* (95% CI) reports unstandardized simple slopes and standardized mean differences (Cohen's *d*) were calculated using the baseline standard deviation of each outcome. Overall well-being estimates are from the linear mixed effects model, while subscale estimates are from the multivariate mixed effects model. Models were adjusted for age and gender identity.

* $p < .05$, ** $p < .01$, *** $p < .001$

**Table S4**

*Interaction Effects for ELA Moderating Mental Health*

| | Marginal Means | | Simple Slopes | |
|---|---|---|---|---|
| | **Low ELA (-1 *SD*)** | **High ELA (+1 *SD*)** | **Baseline** | **Post-Course** |
| **Overall Mental Distress** | $M\Delta$ = -2.28<br>(-5.72, 1.16)<br>$d$ = 0.09 | $M\Delta$ = -9.11***<br>(-12.58, -5.65)<br>$d$ = 0.38 | $B$ = 0.581***<br>(0.414, 0.748) | $B$ = 0.341***<br>(0.162, 0.520) |
| **Overall Mental Well-Being** | $M\Delta$ = 2.68**<br>(0.65, 4.71)<br>$d$ = 0.22 | $M\Delta$ = 6.32***<br>(4.30, 8.34)<br>$d$ = 0.52 | $B$ = -0.264***<br>(-0.370, -0.158) | $B$ = -0.136*<br>(-0.250, -0.022) |

*Note.* Marginal means showing pre-to-post changes $M\Delta$ (95% CI) in mental distress and well-being at low (-1 *SD*) versus high (+1 *SD*) ELA. $B$ (95% CI) reports unstandardized simple slopes and standardized mean differences (Cohen's $d$) were calculated using the baseline standard deviation of each outcome. Estimates are from the multivariate mixed effects model, specifying overall scores for mental distress and well-being as correlated outcomes.

* $p < .05$, ** $p < .01$, *** $p < .001$

**Figure S1**

*Bivariate Correlations Across Study Variables*

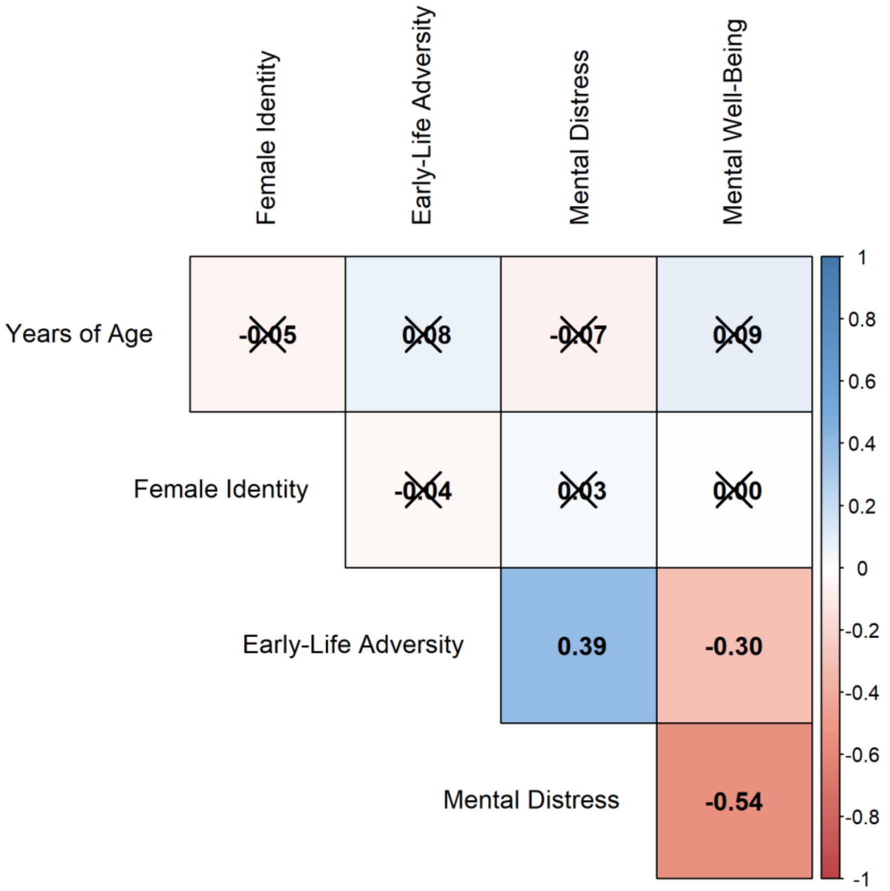


*Note.* Bivariate (parametric) correlations across study variables at baseline. Correlations with female identity (dummy coded: non-female = 0, female = 1) are point-biserial. Diagonal crosses are superimposed over non-significant correlations ($p > .05$) .

**Figure S2**

*Bivariate Correlations Across ELA Types*

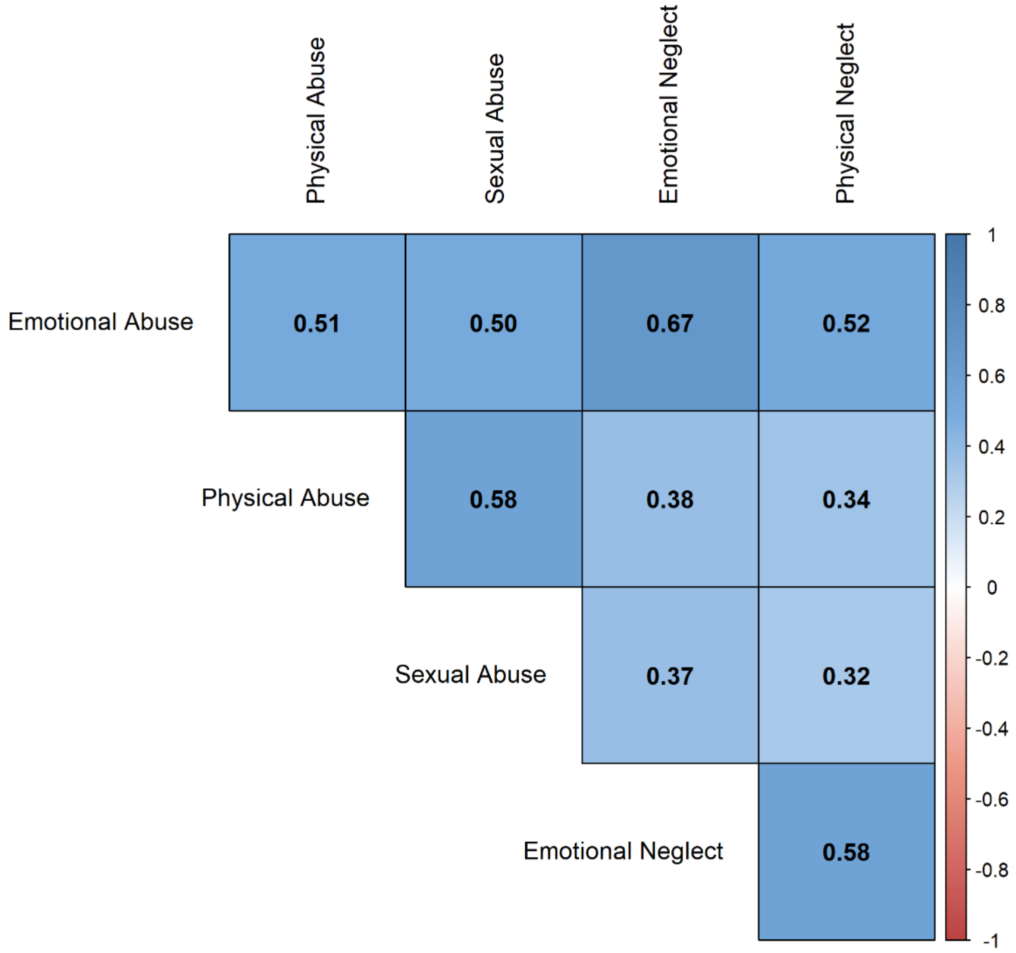


*Note.* Bivariate (parametric) correlations across the different subdomains of ELA. All correlations were significant ($p < .05$).
